# Admissions to a large tertiary care hospital and Omicron BA.1 and BA.2 SARS-CoV-2 PCR positivity: primary, contributing, or incidental COVID-19

**DOI:** 10.1101/2022.04.12.22273760

**Authors:** Anne F. Voor in ’t holt, Cynthia P. Haanappel, Janette Rahamat – Langendoen, Richard Molenkamp, Els van Nood, Leon M. van den Toorn, Robin P. Peeters, Annemarie M.C. van Rossum, Juliëtte A. Severin

**Affiliations:** Department of Medical Microbiology and Infectious Diseases, Erasmus MC University Medical Center Rotterdam, The Netherlands; Department of Viroscience, Unit Clinical Virology, Erasmus MC University Medical Center Rotterdam, The Netherlands; Department of Pulmonology, Erasmus MC University Medical Center Rotterdam, The Netherlands; Department of Internal Medicine, Erasmus MC University Medical Center Rotterdam, The Netherlands; Department of Pediatrics, Division of Infectious Diseases and Immunology, Erasmus MC-Sophia Children’s Hospital University Medical Center Rotterdam, The Netherlands

**Keywords:** COVID-19, patient admission, SARS-CoV-2, epidemiology, Omicron

## Abstract

SARS-CoV-2 Omicron variants BA.1 and BA.2 seem to show reduced clinical severity. We classified 172 COVID-19 Omicron patient admissions. 66.2% of patients were admitted with primary or admission-contributing COVID-19. We therefore must be careful to base healthcare and public health decisions on the total number of hospitalized COVID-19 patients alone.

## Introduction

Monitoring national hospitalization rates for COVID-19 has been essential throughout the pandemic to guide public health decision-making, to evaluate vaccine efficacy, and to implement a wide range of other medical interventions. However, with the rapid worldwide spread of the SARS-CoV-2 Omicron variant of concern and increasing immunity against SARS-CoV-2, interpreting the true impact of these hospitalization rates has been complicated (1, 2).

Signs of reduced clinical severity of Omicron compared to previous variants appeared quickly (3-8), with unvaccinated patients still at the highest risk of substantial morbidity and mortality (9). Omicron variants have been divided into four distinct sublineages: BA.1, BA.1.1, BA.2, and BA.3. Within the Netherlands, BA2 is currently identified as the predominant strain Given the different backgrounds (*e.g*., seroprevalence, vaccination rates) of countries reporting reduced clinical severity of the Omicron variant, the monitoring of national hospitalization rates is essential to predict the overload of healthcare and leads the way for measures in the community.

Because of the rapid spread of Omicron and the subsequent high incidence of infection in the population, not all SARS-CoV-2 positive patients admitted to the hospital have been admitted solely because of COVID-19. It is important to distinguish between admissions that are due to primary COVID-19, admissions where COVID-19 contributes but is not the only reason (admission-contributing COVID-19), or admissions where COVID-19 is not contributing to the reason of admission (incidental COVID-19). Therefore, we aimed to assess and classify the cause of hospitalization for COVID-19 patients identified with the Omicron variant within our hospital in order to provide more insight into the clinical severity of Omicron and the COVID-19 hospital burden.

## Methods

This study was performed in the Erasmus MC University Medical Center in Rotterdam, The Netherlands (Erasmus MC). The Erasmus MC is a large tertiary care hospital with 1125 beds, with a total of 121 beds at the intensive care unit (ICU). A retrospective analysis was performed on all patients identified with the SARS-CoV-2 Omicron variant between 23 December 2021 and 27 February 2022. In this period, the testing strategy of the hospital was based on symptoms. SARS-CoV-2 infection was identified by real-time transcription-mediated amplification with the Aptima® SARS-CoV-2 assay using the Panther system (Hologic, Malborough, USA), or by the Xpert® Xpress SARS-CoV2 assay on a GeneXpert® system (Cepheid®, Sunnyvale, USA). SARS-CoV2 positive samples were further characterized by variant-specific PCR using VirSNiP (TIBmolbiol, Berlin, Germany) assays targeting S371-S373 and K417 as proxy for the Omicron variant. While detection of K417N was considered indicative for Omicron, S371L-S373P was considered indicative for BA.1 and S371F-S373P for BA.2.

SARS-CoV-2 Omicron variant positive patients at admission were divided in the following categories: 1) patients who were hospitalized for more than 24 hours within 7 days of their first positive SARS-CoV-2 PCR, 2) patients who were hospitalized for less than 24 hours within 7 days of their first positive SARS-CoV-2 PCR, and 3) patients with visits only to the Erasmus MC outpatient clinic. Data was collected from electronic health records (EHR) and included basic patient characteristics such as age and sex, and information about country of birth, vaccination status, admission information (reason of admission, wards of admission, admission date and discharge date), and oxygen therapy during admission.

Patients with a positive SARS-CoV-2 PCR upon clinical admission or during clinical admission were classified based on modified definitions developed by the National Intensive Care Evaluation (NICE) Foundation (10). Classification 1) primary COVID-19; COVID-19 is the main cause for hospitalization; 1A) the patient is hospitalized due to COVID-19 symptoms and is receiving medical treatment for these symptoms, 1B) the patient is hospitalized due to COVID-19 symptoms, does not receive medical treatment, but is admitted for observation due to the underlying disease. Classification 2) admission-contributing COVID-19; COVID-19 is one of the causes for hospitalization; 2A) the patient is admitted for another medical cause but also has COVID-19 symptoms and is receiving medical treatment for these symptoms, 2B) dysregulation of underlying disease due to COVID-19 (*e.g*., sickle cell crisis provoked by SARS-CoV-2 without respiratory involvement). Classification 3) incidental COVID-19; COVID-19 is not the cause for hospitalization, the patient does not have any or only mild COVID-19 symptoms, and does not receive any medical treatment for these symptoms. Classification 4) Indeterminate; it is unknown whether the cause of hospitalization is related to COVID-19 symptoms. Classifications were performed by two epidemiologists through evaluation of the patient’s history in the EHR. If the classification was unclear patients were assessed separately by two independent epidemiologists before reaching agreement of classification.

This study was approved by the medical ethical research committee of the Erasmus MC (MEC-2021-0845-A-0002), and was not subject to the Medical Research Involving Human Subjects Act.

## Results

A total of 402 patients were identified with the Omicron variant of SARS-CoV-2 within the Erasmus MC. A total of 333 adult patients were identified with the Omicron variant, of whom 287 patients with variant BA.1 (86.1%), 28 patients with variant BA.2 (8.4%), and 18 patients with Omicron variant for which further distinction in BA.1 or BA.2 could not be made due to low viral load in the sample (5.4%). Of patients with BA.1, 39.4% had a clinical admission of more than 24 hours, 9.8% had a clinical admission of fewer than 24 hours, and 50.9% had an outpatient visit only. For adult patients with BA.2, 50% were clinically admitted to the hospital and 50% had an outpatient visit. Ninety-six pediatric patients were identified with the Omicron variant. BA.1 was identified in 57 children (82.6%), BA.2 in 16 children (15.9%), and Omicron without further distinction in BA.1 or BA.2 in 1 child (1.4%). Of children identified with BA.1, 26.3% were clinically admitted for more than 24 hours, while 1.8% had a clinical admission of fewer than 24 hours, and 71.9% had an outpatient visit only. For children with BA.2, 36.4% had a clinical admission of more than 24 hours and 63.6% only had an outpatient visit.

One hundred seventy-two patients were hospitalized for more than 24 hours at the Erasmus MC and were identified with the Omicron variant of SARS-CoV-2 at admission or during admission (Table 1). Out of these 172 patients, 143 patients were identified with Omicron variant BA.1 (83.1%), 19 patients with BA.2 (11.0%), and 10 patients with Omicron without further distinction in BA.1 or BA.2 (5.8%) (Table 1).

**Table 1.** Characteristics of included hospitalized adult and pediatric COVID-19 patients for each classification.

|  | Total | Clas1:<br>Primary<br>COVID-19 | Clas2:<br>Contributing<br>COVID-19 | Clas3:<br>Incidental<br>COVID-19 | Clas4:<br>Indeterminate<br>COVID-19 |
| --- | --- | --- | --- | --- | --- |
| **Adult patients (%)** | 151 (100) | 68 (45.0) | 32 (21.2) | 46 (30.5) | 5 (3.3) |
| Median age (range) | 56 (18-90) | 61 (18-90) | 51 (18-82) | 55 (22-90) | 23 (22-49) |
| Male gender (%) | 82 (54.3) | 35 (51.5) | 20 (62.5) | 25 (54.3) | 2 (40) |
| Country of birth |  |  |  |  |  |
| The Netherlands | 90 (60.4)<sup>2</sup> | 44 (65.7)<sup>3</sup> | 18 (56.3) | 26 (57.8)<sup>3</sup> | 2 (40) |
| Other | 59 (39.6)<sup>2</sup> | 23 (34.3)<sup>3</sup> | 14 (43.8) | 19 (42.2)<sup>3</sup> | 3 (60) |
| Solid organ recipient (%) | 23 (15.2) | 16 (23.5) | 6 (18.8) | 1 (2.2) | na |
| Lung (%) | 9 (6.0) | 8 (11.8) | 1 (3.1) | 0 (0) | na |
| Kidney (%) | 13 (8.6) | 8 (11.8) | 4 (12.5) | 1 (2.2) | na |
| 28-day in-hospital mortality (%) | 12 (7.9) | 7 (10.3) | 1 (3.1) | 4 (8.7) | 0 (0) |
| Vaccinated<sup>1</sup> | 55 (53.9)<sup>4</sup> | 37 (61.7)<sup>5</sup> | 7 (30.4)<sup>6</sup> | 11 (57.9)<sup>11</sup> | na |
| Received booster vaccination | 33 (34.7)<sup>7</sup> | 25 (43.1)<sup>8</sup> | 3 (14.3)<sup>9</sup> | 5 (31.3)<sup>12</sup> | na |
| BA.1 lineage (%) | 127 (84.1) | 52 (76.5) | 27 (84.4) | 43 (93.5) | 5 (100) |
| BA.2 lineage (%) | 14 (9.3) | 9 (13.2) | 2 (6.3) | 3 (6.5) | 0 (0) |
| Oxygen therapy during admission | 74 (50.7)<sup>10</sup> | 56 (82.4) | 13 (40.6) | 5 (10.9) | na |
| ICU during admission (%) | 13 (8.6) | 6 (8.8) | 1 (3.1) | 6 (13.0) | 0 (0) |
| ICU as admission department (%) | 10 (6.6) | 5 (7.4) | 0 (0) | 5 (10.9) | 0 (0) |
| **Pediatric patients (%)** | 21 (100) | 4 (19.0) | 6 (28.6) | 8 (38.1) | 3 (14.3) |
| Median age (range) | 3 (0-17) | 4 (0-8) | 3 (0-13) | 2 (0-7) | 15 (12-17) |
| Male gender (%) | 12 (57.1) | 2 (50) | 5 (83.3) | 4 (50) | 1 (33.3) |
| Country of birth |  |  |  |  |  |
| The Netherlands | 19 | 4 (100) | 5 (83.3) | 7 (87.5) | 3 (100) |
| Other | 2 | 0 (0) | 1 (16.7) | 1 (12.5) | 0 (0) |
| 28-day in-hospital mortality (%) | 0 (0) | 0 (0) | 0 (0) | 0 (0) | 0 (0) |
| BA.1 lineage (%) | 16 (76.2) | 2 (50) | 5 (83.3) | 7 (87.5) | 2 (66.7) |
| BA.2 lineage (%) | 5 (23.8) | 2 (50) | 1 (16.7) | 1 (12.5) | 1 (33.3) |
| Oxygen therapy during admission (%) | 8 (38.1) | 4 (100) | 3 (50) | 1 (12.5) | na |
| ICU during admission (%) | 6 (28.6) | 1 (25) | 3 (50) | 2 (25) | 0 (0) |
| ICU as admission department (%) | 6 (28.6) | 1 (25) | 3 (50) | 2 (25) | 0 (0) |

Abbreviations: ICU; intensive care unit, na; not available, clas; classification.
<sup>1</sup>Received 2 COVID-19 vaccinations (Pfizer/BioNTech, Moderna or AstraZeneca) or 1 vaccination (Johnson & Johnson's Janssen), <sup>2</sup> for 2 patients this was unknown, <sup>3</sup> for 1 patient this was unknown, <sup>4</sup> for 49 patients this was unknown, <sup>5</sup> for 8 patients this was unknown, <sup>6</sup> for 9 patients this was unknown, <sup>7</sup> for 56 patients this was unknown, <sup>8</sup> for 10 patients this was unknown, <sup>9</sup> for 11 patients this was unknown, <sup>10</sup> for 5 patients this was unknown, <sup>11</sup> for 27 patients this was unknown, <sup>12</sup> for 30 patients this was unknown.

## Discussion

This study assessed and classified the cause of hospitalization of 172 SARS-CoV-2 positive patients. We observed that less than 50% of adult Erasmus MC patients identified with the Omicron variant were clinically admitted for more than 24 hours. Of these, 45% were primary COVID-19 cases, 21% were contributing, and 31% were incidental. Primary COVID-19 patients were older, had a higher 28-day in-hospital mortality rate, and showed more BA.2. However, they had higher COVID-19 vaccination and booster rates compared to incidental COVID-19 cases. Additionally, primary COVID-19 and admission-contributing COVID-19 were more frequently observed among transplant recipients. These groups differ thereby from patients with incidental COVID-19 Omicron patients. This is in line with the study conducted by Sun *et al*. (11).

Studies in pediatric patients show that the number of admitted pediatric patients with the Omicron variant is higher compared to previous variants (12). In this study, we did not register any in-hospital mortality among pediatric patients and small numbers of ICU admissions suggesting no increased clinical severity, even though incidence among children was high in the community.

Initial studies on patients with the Omicron variant mainly assessed the change in clinical severity of hospitalized COVID-19 patients. However, these studies did not differentiate between primary and incidental COVID-19, hereby providing a general conclusion for all patients, while inherent differences are to be expected between patient populations (3, 4)Studies have reported between 17.6% and 19.7% of hospitalized COVID-19 patients with the Omicron variant receiving oxygen therapy and an in-hospital mortality between 2.7 and 5.8%, while we registered over 80% of primary COVID-19 patients receiving oxygen therapy and a mortality of 10.3%(4, 6) This could be attributed to the larger proportion of immunocompromised patients among primary COVID-19 admissions.

We suggest assessing the risk of severe disease caused by SARS-CoV-2 differently for primary COVID-19 compared to incidental COVID-19 patients. These data are of interest to clinicians but also to stakeholders such as hospital managers, infection prevention and control (IPC) practitioners, public health professionals, and the general public. Weighing these patient groups separately in terms of the attributed burden for healthcare could have implications for public health and healthcare decision-making. This could include maintaining non-pharmaceutical interventions such as masks for patients at risk of becoming a primary COVID-19 patient (11).

Although both primary and incidental COVID-19 hospitalizations have implications for workload and isolation capacity, incidental COVID-19 in patients generally interferes less with continuity of care. Counting patients with incidental COVID-19 as COVID-19 admissions therefore gives a skewed image of hospital workload and the COVID-19 burden we are currently dealing with. Therefore, one should be careful to base healthcare and public health decisions, in the evolving landscape of COVID-19 on the total number of hospitalized COVID-19 patients alone.

## Data Availability

All data produced in the present study are available upon reasonable request to the authors.

